# Evaluation of Depression, Anxiety and Sleep Quality in the Brazilian Population During Social Isolation Due to the New Coronavirus (SARS-CoV-2) pandemic: the DEGAS-CoV Study

**DOI:** 10.1101/2021.02.02.21250825

**Authors:** Paulo Afonso Mei, Amanda Sasse, Ana Lara Navarrete Fernandez, Bárbara Neiva Perri, Breno Alexander Bispo, Cintia Zonta Baptista, Fábio Soares Néspoli, Gabriela Sakita Munhos, Giovanni Giuliani Verghetti, Giselly Brito Santana, Guilherme Barbosa de Almeida Oliveira Martins, Jennifer Pereira da Rocha, Jessyca Rosa Lopes Mendonça, Julia Patel Lebl, Laís Grabner Ruivo, Laura Loeb, Marielly Isepon, Marina Joseane Pachecco, Paloma Ricciardi de Castro, Paola Ricciardi de Castro, Rafaela Dotta Brustolin, Taysa Ma. P. Gonçalves G. da Silva, Valdemiro Da Rolt, Victoria Gomes Andreata, Amilton dos Santos, Tânia Aparecida Marchiori Oliveira Cardoso

## Abstract

**Introduction:** The new coronavirus infection (COVID-19) has caused distress and repercussions in mental and physical health of individuals. Depression, anxiety and worsening of sleep quality have been reported in several recent articles that surveyed populations all over the globe. Our work meant to access, through a cross-sectional study, these disorders in the Brazilian population, through the application of an online questionnaire conducted on the second trimester of 2020.

**Materials and Methods:** We applied an online questionnaire, filled with questions regarding social, economic, financial, educational and health status, as well as questions from the Hospital Anxiety and Depression Scale (HAD), and from the Pittsburgh Sleep Quality Index (PSQI).

**Results:** We collected 2,695 valid answers, from April 24^th^ to May 31^st^, 2020. Age ranged from 18 to 79 years, mean of 31.3. Women were 76.3%, men 23.7%. Symptoms of Anxiety were found in 56.5%, of depression in 46.1%, and of bad sleep in 49.2%. Some groups were more prone than others to one or more of those conditions, such as: younger people, women, mestizos, Northeasterners, people with lesser years of education, of lower income or whose income dropped significantly during the pandemic, caregivers, students, sedentary or people practicing less physical activity, people who followed more hours of news of COVID-19 and those less engaged in social and instrumental activities.

**Conclusion:** anxiety, depression and bad sleep quality were significantly high in our survey. Mental and sleep health is heterogeneously affected among individuals, depending on social, economic, financial, educational and health status.

**HIGHLIGHTS**

- An online survey (DEGAS-CoV) was conducted between April 30^th^ and May 31^st^, 2020, with people living in Brazil, aged 18 or more. The study obtained 2,695 valid answers.
- Rates of possible anxiety, possible depression and bad sleep quality were 56.5%, 46.1% and 49.2%, respectively. Rates are similar to another Brazilian survey, with 45,161 participants, conducted in a similar time window.
- Were more prone to mental and/or sleep conditions: younger participants, women, mestizos, unemployed, students, people with less years of education, people with lower income or with considerable drops of income during the virus outbreak, caregivers, people who followed more news of COVID-19, people less engaged in social and instrumental activities, smokers, sedentary or those who practiced less physical activity, and people who had symptoms suspected (confirmed or not) of SARS-CoV-2 infection.
- Alcohol drinkers were slightly less likely to be possibly depressed. That finding needs more clarification and may be due to confounders.

## I. Introduction

Since December 2019, after the first report in China (1), the new strain of Coronavirus, prompter of a severe acute respiratory syndrome (SARS-CoV-2), designated as COVID-19 by the World Health Organization (2), has caused and unprecedent, worldwide distress. Public health measures are taken in many cases, in an improvised fashion and within a blink of an eye. In short intervals social restrictions might be lifted, just be implemented again later, due to the erratic behavior of the virus and oscillations in its transmission ratio and also the evidence of secondary waves of outbreak (3).

In an unsteady scenario of a deadly virus outbreak, with massive misinformation, added to the negative effect of social restrictions, a rise in the prevalence of mental disorders, such as depression and anxiety, and disruptions in the normal, physiological sleep are widely expect to occur, with possible short- and long-term consequences (4) (5). Previous studies with data from past epidemic disorders, such as in H1N1 influenza in 2009 (6), SARS-CoV in 2002 and Middle East Respiratory Syndrome (MERS) in 2012 (7) attest this fact.

One of the first to access mental and sleep issues during the outbreak of COVID-19, Huang and Zhao applied an online questionnaire in 7,236 people living in China, finding anxiety, depression and bad sleep ratios of 35%, 20% and 18%, respectively (8).

Several other studies conducted in countries in all continents, showed an increase of anxiety, depression, bad sleep quality and/or stress as well (8) (9) (10) (11) (12) (13) (14) (15) (16) (17). One study from the United Kingdom (12) found that higher levels of anxiety were associated with more somatic symptoms, like gastrointestinal disturbances or fatigue, hence impoverishing life quality in such cases. In Brazil, one study accessed mental and sleep complaints during COVID-19 in more than 45,000 people, surveyed through an online form, and found 52.6% of possible anxiety, 41.4% of possible depression and 43.5% of bad sleepers (18).

This study intended to measure the overall prevalence of anxiety and depression symptoms, and bad sleep quality in the second trimester of 2020, among people living in Brazil, while moderate to severe social restrictions were in place, and compare rates between different groups, according to social, financial, educational, health and economic status.

## II. Materials and Methods

### II-A. Ethical aspects, inclusion criteria and study design

We conducted a cross-sectional study, called DEGAS-CoV, an acronym for “Depression, General Anxiety and Sleep Disorders during COVID-19”. The study was based on an online survey from April 30^th^ to May 31^st^ 2020, when measures establishing some degree of social distancing – governmental advice or imposition of restrictions like closure of commerce, surveillance of circulation of cars and pedestrians, and even lock-down in more critical areas were in place, in Brazil.

Before commencing, ethical approval was granted by the São Leopoldo Mandic Medical College Ethical Committee. Dissemination and recruitment of participants was made entirely online, through snowball sampling (19), keeping in mind the current scenario of social restrictions. Participants were approached by the researchers either directly, via social medias (i.e., WhatsApp, Facebook and others), or indirectly, when other participants, in their turn, shared the link to the survey with their acquaintances.

Participants were required to be at least 18 years old, and to be currently living in Brazilian soil. No physical or mental conditions/disorders were considered as exclusion criteria. Both people who already contracted or did not contract COVID-19 were included as well.

Before answering, the participant was presented with a consent form and was required to check that he/she had read and agreed with all conditions.

### II-B. Questionnaires Applied

The online form was divided in three questionnaires, and was created using Google Forms (20).

The first questionnaire consisted of interrogations formulated by the researchers, and meant to access geographical, educational, financial, and health status of participants, as well as habitual behaviors, such as social activities (SA) – phone or video calls/conferences and in-person interactions with familiars, friends or other people - and instrumental activities (IA) – care of offspring/parents/relatives, pet sitting, preparing meals, cleaning the house and other inhouse daily activities, shopping, religious/spiritual worship - and time spent watching or reading news. Lastly, patients were required to answer two questions regarding COVID-19. First, if they already contracted COVID-19 and, if not, how afraid they were of contracting – possible answers were “already contracted”, “no or little fear”, “moderate fear”, and “intense fear of contracting”. Secondly, they were asked to grade, in a Likert scale fashion, from 1 to 5, how severely COVID-19 had affected their lives, in general, being 1 “no/very low interference” and 5 “total interference”.

The second questionnaire was composed of the Hospital Anxiety and Depression Scale - HAD (21), consisting of 14 multiple-choice questions, 7 for the evaluation of possible depression and 7 for possible anxiety. The Brazilian Portuguese validated translation (22) was used. The higher the frequency and/or intensity of symptoms, the higher the punctuation. A cut-off of 9 points for each symptom was considered for tagging the responder as a bearer of those conditions.

For the last questionnaire, researchers used a modified version of Brazilian validated translation (23) of the Pittsburgh Sleep Quality Index - PSQI (24), what was called mPSQI. This instrument is designed for the triage of people with sleep of bad quality, regardless of etiology. The PSQI score is calculated by the sum of components (subset of questions), each receiving a determined weight. For most authors, a cut-off score of 5 points is adequate for the identification of bad sleepers (25).

The original version of the PSQI consists of 10 questions, being the first 4 open questions dealing with a free estimate of time of going to bed and getting up, sleep latency in minutes and effective hours of sleep, and the last 6 questions of multiple-choice; the higher the frequency and/or intensity of symptoms, the higher the punctuation. In the original version, the maximum score is 21 points (24).

For the mPSQI, we opted for suppression of the first 4 and also of the 10^th^ question, leading to a maximum possible punctuation of 15 points. For the first 4 questions, we felt that calculation and estimation of time could offer some difficulty to some responders, particularly those with lesser education years, given that no presential support could be offered. Question 10 was not included because, besides not being considered for scoring, it required the participant to have a partner or other roommate to attest snoring, and given the unusual circumstances, a considerate fraction of people enrolled could not have an informant. The cut-off commonly used value of five points for the PSQI was maintained for the mPSQI, as there was no certainty of sensitivity/specificity by adopting a lower cut-off for the mPSQI.

### II.C Data Analysis

Statistical Analysis was conducted in the Statistical Package for Social Sciences (SPSS) suite (26). Categorical variables were compared by Chi-Square test, with adopted significance level of 95%. For contingency tables with more than 1 degree of freedom, a Post-Hoc analysis was conducted, being the p value (α) modified by Bonferroni correction (27), for a contingency table of *i* lines and *j* columns, α = 0.05 / (*i* x *j*). Odd ratios were calculated by univariate logistic regression. When needed, correlation was made by Spearman 2-tailed Correlation index (ρ), since data was non-parametric.

## III. Results

### III-A. Exploratory analysis

The questionnaire received a total of 2,839 inputs. After exclusions, either due to duplicity (last answer was validated) or to incoherent responses, a total of 2,695 forms remained. Mean age was of 31.3 years of age; 2,052 (76.3%) were women and 636 (23.7%) men; most auto-declared themselves as whites, followed by mestizos, blacks, Asians and indigenous. Epidemiological data of participants are further detailed in **Table 1**, ahead.

**Table 1.** Exploratory analysis of different aspects of responders; for each category total valid was 2,695, unless specified

| Condition | N | % | subsets, when suitable | N (%) |
| --- | --- | --- | --- | --- |
| **A - DEMOGRAPHICS** |  |  |  |  |
| All responders | 2,695 | 100 |  |  |
| AGE |  |  | Limits |  |
| Mean (std deviation = 0.236) | 31.27 | -- | Low | 18 |
| Median | 27 | -- | High | 19 |
| SEX (Total valid = 2,688) |  |  |  |  |
| Women (cis or trans) | 2,052 | 76.3 |  |  |
| Men (cis or trans) | 636 | 23.7 |  |  |
| Race - auto declaration (Total valid = 2,652) |  |  |  |  |
| White | 1,964 | 74.1 |  |  |
| Mestizos | 515 | 19.4 |  |  |
| Black | 116 | 4.4 |  |  |
| Asian | 48 | 1.8 |  |  |
| Indigenous | 9 | 0.3 |  |  |
| # people living together during the pandemic |  |  |  |  |
| Responder lives alone | 184 | 6.8 |  |  |
| Lives with more people | 2,511 | 93.2 |  |  |
| **B – EDUCATION/OCCUPATION** |  |  |  |  |
| Schooling |  |  |  |  |
| Incomplete Elementary/Middle | 5 | 0.2 |  |  |
| Completed Middle, never started High School | 29 | 1.1 |  |  |
| Incomplete High School | 17 | 0.6 |  |  |
| Completed High School (may have started University but did not complete) | 1182 | 43.9 |  |  |
| Completed graduation | 726 | 26.9 |  |  |
| Completed postgraduation | 736 | 27.3 |  |  |
| Is the person currently a student? |  |  |  |  |
| Yes, person is currently a student | 1,162 | 43.1 |  |  |
| No, person is not currently a student | 1,533 | 56.9 |  |  |
| Main Occupation Type, General |  |  |  |  |
| Unemployed | 125 | 4.6 |  |  |
| Freelancer | 495 | 18.4 |  |  |
| Private sector | 762 | 28.3 |  |  |
| Public employee | 48 | 1.8 |  |  |
| Student | 1,162 | 43.1 |  |  |
| Retired/Receiving pension | 103 | 3.8 |  |  |
| Occupation in Health management |  |  |  |  |
| Non-health worker | 2,005 | 74.4 |  |  |
| Health worker/student, not involved in care of COVID | 519 | 19.3 |  |  |
| Health worker/student, involved in care of COVID patients | 170 | 6.3 |  |  |
| Caregiving |  |  |  |  |
| Not a caregiver | 2,590 | 96.1 |  |  |
| Caregiver | 105 | 3.9 | Caregiver – one person | 85 (3.2) |
|  |  |  | Caregiver – two people | 14 (0.5) |
|  |  |  | Caregiver – more than 2 p. | 6 (0.2) |
| **C – FINANTIAL STATUS** |  |  |  |  |
| Family Income (Total valid = 2,388) |  |  |  |  |
| Less than R\$ 1,200/month | 102 | 4.3 | | |
| R\$ 1,200 – 3,000/month | 498 | 20.8 | | |
| R\$ 3,001 – 10,000/month | 1,021 | 42.7 | | |
| Above R\$ 10,000/month | 768 | 32.1 | | |
| Change of family income |  |  |  |  |
| Income did not drop | 993 | 36.8 | Income has improved | 35 (1.3) |
|  |  |  | Income hasn't changed | 958 (35.5) |
| Income dropped | 1,702 | 63.2 | Up to 25% drop | 863 (32.0) |
|  |  |  | 25 – 50% drop | 567 (21.0) |
|  |  |  | More than 50% drop | 272 (10.1) |
| Provider of family income |  |  |  |  |
| Responder is the main provider | 727 | 27.0 | Is the sole provider | 283 (10.5) |
|  |  |  | Other members contribute with a lesser proportion | 443 (16.4) |
| Responder is not the main provider | 1,968 | 73.0 | Responder contributes with some amount | 858 (31.8) |
|  |  |  | Responder does not contribute at all | 1,111 (41.2) |
| **D – PHYSICAL CONDITIONS, COMORBIDITIES, HABITS AND BEHAVIORS** |  |  |  |  |
| BMI (Total valid = 2,691) |  |  |  |  |
| Normal | 1,500 | 55.7 |  |  |
| Overweight | 771 | 28.6 |  |  |
| Obese | 421 | 15.6 |  |  |
| Comorbidities or other health conditions |  |  |  |  |
| No comorbidities | 1959 | 72.7 |  |  |
| With comorbidity(s)/condition | 736 |  | 1 comorb., pregnancy excluded | 574 |
|  |  |  | 2 comorb., pregnancy excluded | 120 |
|  |  |  | 3 comorb., pregnancy excluded | 25 |
|  |  |  | 4 + comorb., pregnancy excluded | 10 |
| Smoker |  |  |  |  |
| No | 2,507 | 93.1 |  |  |
| Yes | 188 | 6.9 | Up to 10 cigarettes/day | 128 (4.7) |
|  |  |  | More than 10 cig./day | 60 (2.2) |
| Alcohol intake |  |  |  |  |
| No | 1,523 | 56.5 |  |  |
| Yes | 1,172 | 43.5 |  |  |
| Symptoms of COVID-19 |  |  |  |  |
| No symptoms | 2,296 | 85.2 |  |  |
| Had symptoms | 399 | 14.8 | COVID-19 test was negative | 76 (2.8) |
|  |  |  | Still waiting results/did not test for COVID-19 | 295 (10.9) |
|  |  |  | COVID-19 test was positive | 28 (1) |
| Fear of contracting COVID-19 |  |  |  |  |
| Has already contracted | 28 | 1.0 |  |  |
| Did not contract to the date | 2,667 | 99.0 | Not or little worried about | 433 (16.8) |
|  |  |  | Moderately worried | 1,405 (52.1) |
|  |  |  | Very much worried | 809 (30) |
| Time spent gathering news about COVID-19 on TV and other media |  |  |  |  |
| Does not follow news | 134 | 5 |  |  |
| Follows the news |  |  | Up to one hour/day | 1,515 (56.2) |
|  |  |  | 1-3 hours/day | 742 (27.5) |
|  |  |  | More than 3 hours/day | 304 (11.3) |
| Practice of Exercises |  |  |  |  |
| Did not practice before the pandemic | 960 | 35.6 | Remains sedentary | 735 (27.3) |
|  |  |  | Started practicing | 225 (8.3) |
| Practiced before the pandemic | 1,735 | 64.4 | Is not practicing now | 722 (26.8) |
|  |  |  | Still practices, less than 3x/w | 419 (15.5) |
|  |  |  | Still practices, 3+ times/w | 594 (22.0) |
| Time preferred for practice of exercises - for those currently practicing (Total valid = 1,487) |  |  |  |  |
| Prefers morning | 380 | 25.6 |  |  |
| Prefers afternoon | 361 | 24.3 |  |  |
| Prefers evening | 415 | 27.9 |  |  |
| Varies/no preference | 331 | 22.3 |  |  |
| Time spent on social activities (SA) |  |  |  |  |
| Less than before the pandemic | 512 | 19.0 | Much inferior | 266 (9.9) |
|  |  |  | Somewhat inferior | 246 (9.1) |
| Equal or more than before the pandemic | 2,183 | 81.0 | Equal | 675 (25.0) |
|  |  |  | Somewhat superior | 819 (30.4) |
|  |  |  | Much superior | 689 (25.6) |
| Time spent in instrumental activities (IA) |  |  |  |  |
| Less than before the pandemic | 260 | 9.6 | Much inferior | 122 (4.5) |
|  |  |  | Somewhat inferior | 138 (5.1) |
| Equal or more than before the pandemic | 2,435 | 90.4 | Equal | 620 (23.0) |
|  |  |  | Somewhat superior | 860 (31.9) |
|  |  |  | Much superior | 955 (25.4) |

### III-B Prevalence of Possible Anxiety, Depression and Bad Sleep Quality and Statistical Analysis

Results from the statistical analysis are displayed in **Tables 2a, 2b, 2c, 2d, 3 and 4**.

**Table 2a.** Statistical analysis of different groups, by demographical characteristics (α - adjusted p after Bonferroni correction, * - p < α ; ** - p < 0.001 † - 100% for each subset, NS – chi square test was non-significant, hence post-hoc analysis was not performed in this category)

| Condition | N | % | subsets, when suitable | α | Anxious<br>N (%) | p | Depressed N<br>(%) | p | Bad Sleeper<br>N (%) | p |
| --- | --- | --- | --- | --- | --- | --- | --- | --- | --- | --- |
| Total population | 2,695 | 100 |  | -- | 1,523 (56.5) | -- | 1243 (46.1) | -- | 1,326 (49.2) | -- |
| DEMOGRAPHICS |  |  |  |  |  |  |  |  |  |  |
| Age in years |  |  |  |  |  |  |  |  |  |  |
| 1sr quartile (18-22) | 793 | 100 |  | .006 | 513 (64.7) | <.001** | 418 (52.7) | <.001** | 423 (53.3) | .005* |
| 2<sup>nd</sup> quartile (23-27) | 589 | 100 |  |  | 356 (60.4) | .278 | 282 (47.9) | .317 | 289 (49.1) | .920 |
| 3<sup>rd</sup> quartile (28-38) | 678 | 100 |  |  | 388 (57.2) | .689 | 317 (46.8) | .689 | 328 (48.4) | .617 |
| 4<sup>th</sup> quartile (39-79) | 635 | 100 |  |  | 266 (51.9) | <.001** | 226 (35.6) | <.001** | 286 (45.0) | .016 |
| Gender |  |  |  |  |  |  |  |  |  |  |
| Female | 2,052 | 100 |  | .05 | 1,252 | <.001** | 1,011 (49.3) | <.001** | 1,059 (51.6) | <.001** |
| Male | 636 | 100 |  |  | 265 |  | 227 (35.7) |  | 260 (40.9) |  |
| Race (auto declared) |  |  |  |  |  |  |  |  |  |  |
| Asian | 48 | 100 |  | .005 | 24 (50) | .37 | 23 (47.9) | .76 | 20 (41.7) | NS |
| Native Brazilian | 9 | 100 |  |  | 7 (77,8) | .19 | 3 (33.3) | .42 | 7 (77.8) |  |
| Mestizos | 515 | 100 |  |  | 321 (62,3) | .003* | 277 (53.8) | < .001** | 252 (48.9) |  |
| Black | 116 | 100 |  |  | 69 (59,5) | .48 | 62 (53.4) | .11 | 54 (46.6) |  |
| White | 1,964 | 100 |  |  | 1,077 (54,8) | .004* | 858 (43.7) | < .001** | 968 (49.3) |  |
| # People living together during the pandemic |  |  |  |  |  |  |  |  |  |  |
| Responder lives alone | 184 | 100 |  | .05 | 93 (50.5) | NS | 76 (41.3) | NS | 93 (50.5) | NS |
| Lives with more people | 2,511 | 100<sup>†</sup> | Total of 2 people |  | 342 (57.2) |  | 281 (47.0) |  | 309 (51.7) |  |
|  |  |  | Total of 3 or 4 people |  | 860 (56.2) |  | 690 (45.1) |  | 735 (48.0) |  |
|  |  |  | Total of 5 or 6 people |  | 202 (58.9) |  | 172 (50.1) |  | 168 (49.0) |  |
|  |  |  | More than 6 pp together |  | 26 (65.0) |  | 24 (60.0) |  | 21 (52.5) |  |

**Table 2b.** Statistical analysis of different groups, by educational and occupational characteristics (α - adjusted p after Bonferroni correction, * - p < α ; ** - p < 0.001 † - 100% for each subset, NS – chi square test was non-significant, hence post-hoc analysis was not performed in this category)

| Condition | N | % | subsets, when suitable | $\alpha$ | Anxious<br>N (%) | p | Depressed N<br>(%) | p | Bad Sleeper<br>N (%) | p |
| --- | --- | --- | --- | --- | --- | --- | --- | --- | --- | --- |
| Total population | 2,695 | 100 |  | -- | 1,523 (56.5) | -- | 1243 (46.1) | -- | 1,326 (49.2) | -- |
| EDUCATION/OCCUPATION |  |  |  |  |  |  |  |  |  |  |
| Schooling |  |  |  |  |  |  |  |  |  |  |
| Incomplete Elementary/Middle |  | 100 |  | .004 | 4 (80.0) | .27 | 3 | .53 | 4 (80.0) | .16 |
| Completed Middle |  | 100 |  |  | 18 (62.1) | .55 | 13 | .89 | 15 (51.7) | .76 |
| Incomplete High School |  | 100 |  |  | 10 (58.8) | .84 | 10 | .29 | 7 (41.2) | .48 |
| Completed High School (may have started<br>graduation but did not complete) |  | 100 |  |  | 741 (62.7) | <.001** | 603 | <.001** | 625 (52.9) | <.001** |
| Completed graduation |  | 100 |  |  | 385 (53.0) | .03 | 310 | .031 | 348 (47.9) | .424 |
| Completed postgraduation |  | 100 |  |  | 365 (49.6) | <.001** | 304 | .002* | 327 (44.4) | .003* |
| Is the person currently a student? |  |  |  |  |  |  |  |  |  |  |
| Student | 1,162 | 100 |  | .05 | 733 (63.1) | <.001** | 597 (51.4) | <.001** | 610 (52.5) | * |
| Non-Student | 1,533 | 100 |  |  | 790 (51.5) |  | 646 (42.1) |  | 716 (46.7) |  |
| Main Occupation Type, General |  |  |  |  |  |  |  |  |  |  |
| Unemployed | 125 | 100<sup>†</sup> | Dismissed > 3 months ago | .004 | 44 (62.0) | .368 | 38 (53.5) | .194 | 38 (53.5) | .484 |
|  |  |  | Dismissed up to 3 months ago |  | 38 (71.7) | .021 | 32 (60.4) | .036 | 31 (58.5) | .162 |
| Freelancer | 495 | 100 |  |  | 230 (46.4) | <.001** | 183 (36.9) | <.001** | 232 (46.8) | .230 |
| Private sector | 762 | 100 |  |  | 408 (53.5) | .045 | 338 (44.4) | .230 | 343 (45.0) | .007 |
| Public employee | 48 | 100 |  |  | 24 (50.0) | .368 | 20 (41.7) | .549 | 19 (39.6) | .194 |
| Student | 1,162 | 100 |  |  | 733 (63.1) | <.001** | 597 (51.4) | <.001** | 610 (52.5) | .003* |
| Retired/Receiving pension | 103 | 100 |  |  | 46 (44.7) | .012 | 35 (34.0) | .012 | 53 (51.5) | .617 |
| Occupation in Health management |  |  |  |  |  |  |  |  |  |  |
| Non-health worker | 2,005 | 100 |  | .008 | 1,141 (56.9) | NS | 943 (47.0) | NS | 1,015 (50.6) | .012 |
| Health worker/student, not involved in care<br>of COVID patients | 519 | 100 |  |  | 94 (55.3) |  | 68 (40.0) |  | 75 (44.1) | .057 |
| Health worker/student, involved in care<br>of COVID patients | 170 | 100 |  |  | 288 (55.5) |  | 232 (44.7) |  | 236 (45.5) | .162 |
| Caregiver |  |  |  |  |  |  |  |  |  |  |
| Provides direct care to 1 or more<br>people | 105 | 100 |  | .05 | 73 (69.5) | .006* | 66 (62.9) | <.001** | 66 (62.9) | .004* |
| Does not provide direct care to<br>anybody | 2,590 | 100 |  |  | 1,450 (56.0) |  | 1177 (45.4) |  | 1,260 (48.6) |  |

**Table 2c.** Statistical analysis of different groups, by finantial characteristics (α - adjusted p after Bonferroni correction, * - p < α ; ** - p < 0.001 † - 100% for each subset, NS – chi square test was non-significant, hence post-hoc analysis was not perform

| Condition | N | % | subsets, when suitable | $\alpha$ | Anxious<br>N (%) | p | Depressed N<br>(%) | p | Bad Sleeper<br>N (%) | p |
| --- | --- | --- | --- | --- | --- | --- | --- | --- | --- | --- |
| Total population | 2,695 | 100 |  | -- | 1,523 (56.5) | -- | 1243 (46.1) | -- | 1,326 (49.2) | -- |
| FINANTIAL STATUS |  |  |  |  |  |  |  |  |  |  |
| Family Income |  |  |  |  |  |  |  |  |  |  |
| Less than R\$ 1,200/mo | 102 | 100 | | .006 | 77 (75.5) | <.001** | 62 (60.8) | .004* | 56 (54.9) | .271 |
| R\$ 1,200 – 3,000/mo | 498 | 100 | | | 320 (64.3) | <.001** | 288 (57.8) | <.001** | 276 (55.4) | .004* |
| R\$ 3,001 – 10,000/mo | 1,021 | 100 | | | 600 (58.8) | .110 | 484 (47.4) | .617 | 513 (50.2) | .689 |
| Above R\$ 10,000/mo | 768 | 100 | | | 361 (47.0) | <.001** | 284 (37.0) | <.001** | 343 (44.7) | <.001** |
| Change in family income |  |  |  |  |  |  |  |  |  |  |
| Income did not drop | 993 | 100† | Income has improved | .005 | 19 (54.3) | .764 | 11 (31.4) | .072 | 14 (40.0) | .271 |
|  |  |  | Income hasn't changed |  | 489 (51.0) | <.001** | 405 (42.3) | .003* | 466 (48.6) | .689 |
| Income dropped | 1,702 | 100† | Up to 25% drop |  | 475 (55.0) | .271 | 386 (44.7) | .317 | 403 (46.7) | .072 |
|  |  |  | 25 – 50% drop |  | 357 (63.0) | <.001** | 287 (50.6) | .016 | 288 (50.8) | .368 |
|  |  |  | More than 50% drop |  | 183 (67.3) | <.001** | 154 (56.6) | <.001** | 155 (57.0) | .007 |
| Provider of family income |  |  |  |  |  |  |  |  |  |  |
| Responder is the main provider | 726 | 100† | Is the sole provider | .006 | 147 (51.9) | .110 | 124 (43.8) | .424 | 141 (49.8) | NS |
|  |  |  | Other members contribute with a lesser proportion |  | 225 (50.8) | .007 | 172 (38.8) | .001* | 218 (49.2) |  |
| Responder is not the main provider | 1969 | 100† | Responder contributes with some amount |  | 476 (55.5) | .484 | 408 (47.6) | .317 | 398 (46.4) |  |
|  |  |  | Responder does not contribute at all |  | 675 (60.8) | <.001** | 539 (48.5) | .360 | 569 (51.2) |  |

**Table 2d.**
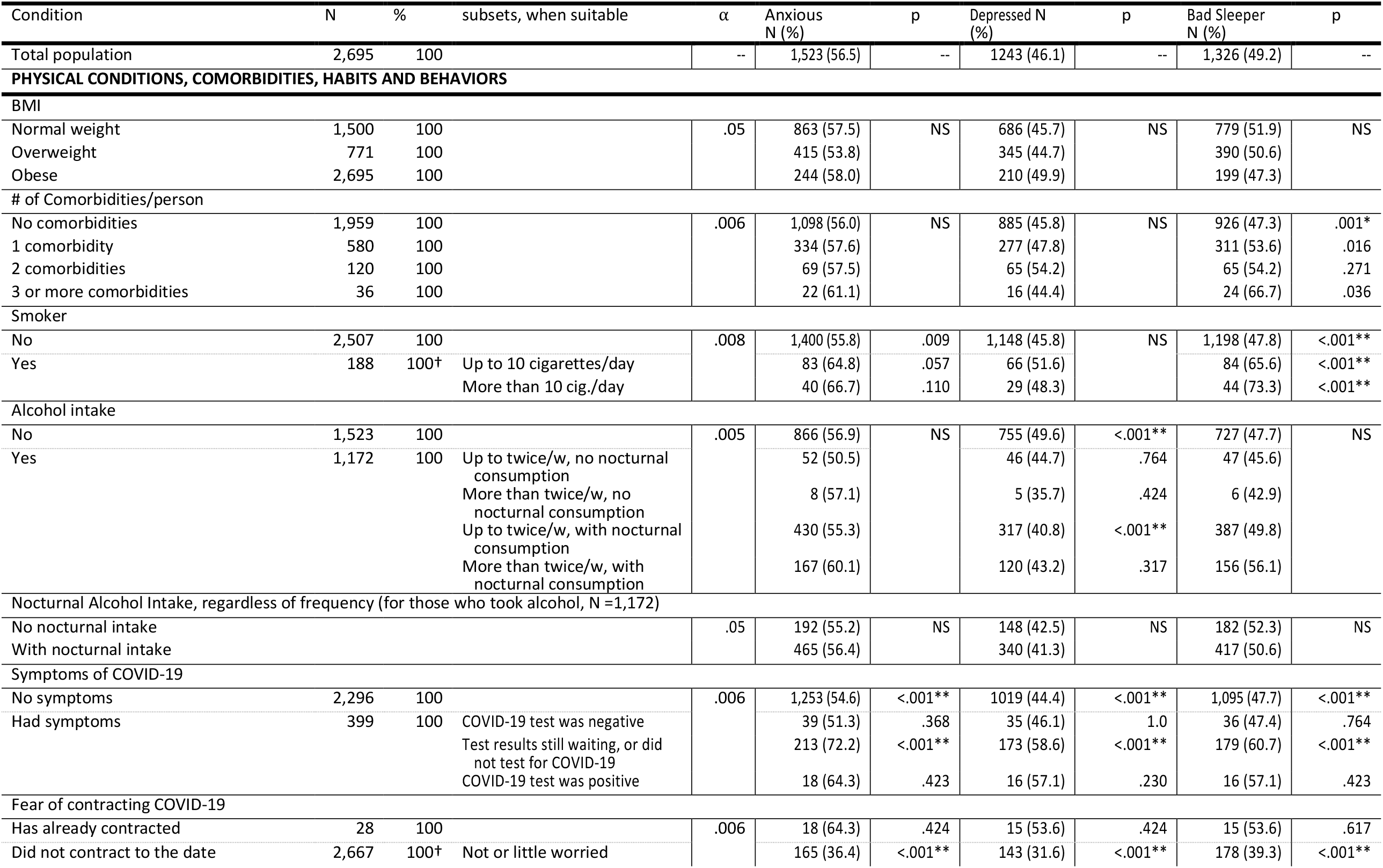
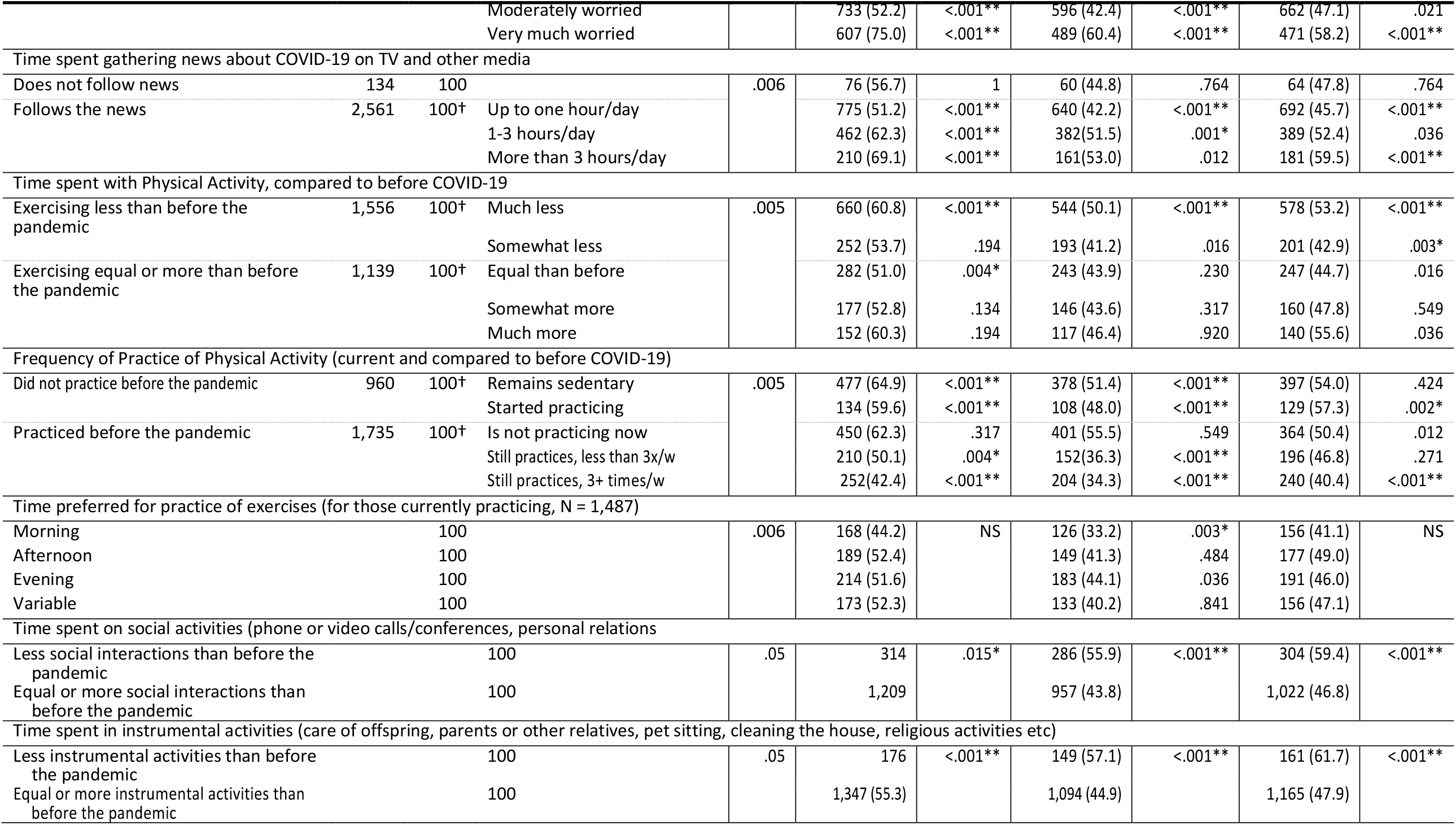
Statistical analysis of different groups, by physical characteristics, comorbidities, habits and behaviors (α - adjusted p after Bonferroni correction, * - p < α ; ** - p < 0.001 † - 100% for each subset, NS – chi square test was non-significant, hence post-hoc analysis was not perform

**Table 3.** Univariate Logistic Regression Odds Ratio (OR) and Confidence Interval (CI) among groups and subsets compared. When more than two subsets were compared, the subset used as reference is stated

| Condition | Anxious<br>OR | CI 95% | Depressed<br>OR | CI 95% | Bad Sleeper<br>OR | CI 95% |
| --- | --- | --- | --- | --- | --- | --- |
| **A - DEMOGRAPHICS** |  |  |  |  |  |  |
| *AGE - Compared to the 4<sup>th</sup> quartile* |  |  |  |  |  |  |
| 1<sup>st</sup> quartile (18-22) | 2.54 | 2.05-3.15 | 2.02 | 1.63-2.50 | 1.40 | 1.13-1.72 |
| 2<sup>nd</sup> quartile (23-27) | 2.12 | 1.69-2.66 | 1.66 | 1.32-2.09 | 1.18 | 0.94-1.47 |
| 3<sup>rd</sup> quartile (28-38) | 1.86 | 1.49-2.31 | 1.59 | 1.27-1.98 | 1.14 | 0.92-1.42 |
| 4<sup>th</sup> quartile (39-79) | 1 |  | 1 |  | 1 |  |
| **SEX** |  |  |  |  |  |  |
| Women (cis or trans) | 2.19 | 1.83-2.63 | 1.75 | 1.46-2.10 | 1.54 | 1.29-1.85 |
| Men (cis or trans) | 1 | -- | 1 | -- | 1 | -- |
| *Race (auto declaration) - Other races compared to "Whites"* |  |  |  |  |  |  |
| White | 1 | -- | 1 | -- | 1 | -- |
| Mestizos | 1.36 | 1.12-1.66 | 1.50 | 1.24-1.82 | 0.99 | 0.81-1.20 |
| Black | 1.21 | 0.83-1.77 | 1.48 | 1.02-2.16 | 0.89 | 0.62-1.30 |
| Asian | 0.82 | 0.46-1.46 | 1.19 | 0.67-2.10 | .74 | 0.41-1.31 |
| Indigenous |  |  |  |  |  |  |
| *# people living together during the pandemic* |  |  |  |  |  |  |
| Responder lives alone | 1 | -- | 1 | -- | 1 | -- |
| Lives with more people | 1.29 | 0.96-1.75 | 1.23 | 0.91-1.67 | 0.94 | 0.70-1.27 |
| **B – EDUCATION/OCCUPATION** |  |  |  |  |  |  |
| *Schooling - All compared to "Completed postgraduation"* |  |  |  |  |  |  |
| Incomplete Elementary/ Middle*** | 4.07 | 0.45-36.55 | 2.13 | 0.35-12.83 | 5.00 | 0.56-44.98 |
| Completed Middle, never started High School | 1.66 | 0.78-3.57 | 1.16 | 0.55-2.44 | 1.34 | 0.64-2.82 |
| Incomplete High School | 1.45 | 0.55-3.86 | 2.03 | 0.76-5.39 | 0.88 | 0.33-2.33 |
| Completed High School (may have started University but did not complete) | 1.71 | 1.42-2.06 | 1.48 | 1.23-1.78 | 1.40 | 1.17-1.69 |
| Completed graduation | 1.15 | 0.94-1.41 | 1.06 | 0.86-1.30 | 1.15 | 0.94-1.42 |
| Completed postgraduation | 1 | -- | 1 | -- | 1 | -- |
| *Is the person currently a student?* |  |  |  |  |  |  |
| Yes, person is currently a student | 1.60 | 1.38-1.88 | 1.45 | 1.25-1.69 | 1.26 | 1.08-1.47 |
| No, person is not currently a student | 1 | -- | 1 | -- | 1 | -- |
| *Unemployed vs Others* |  |  |  |  |  |  |
| Unemployed | 1.53 | 1.05-2.24 | 1.55 | 1.07-2.22 | 1.31 | 0.91-1.89 |
| Employed/Student/Retired | 1 | -- | 1 | -- | 1 | -- |
| *Occupation in Health management* |  |  |  |  |  |  |
| Non-health worker | 1 | -- | 1 | -- | 1 | -- |
| Healthcare worker/student | 0.94 | 0.79-1.12 | 0.87 | 0.73-1.04 | 0.80 | 0.68-0.93 |
| *Caregiving* |  |  |  |  |  |  |
| Not a caregiver | 1 | -- | 1 | -- | 1 | -- |
| Caregiver | 1.79 | 1.18-1.74 | 2.03 | 1.36-3.04 | 1.79 | 1.19-2.67 |
| **C – FINANTIAL STATUS** |  |  |  |  |  |  |
| *Family Income - All compared to "Above R\$ 10,000/month"* | | | | | | |
| Less than R\$ 1,200/month | 3.47 | 2.16-5.57 | 2.64 | 1.73-4.04 | 1.51 | 1.00-2.29 |
| R\$ 1,200 – 3,000/month | 2.03 | 1.61-2.56 | 2.34 | 1.86-2.94 | 1.54 | 1.23-1.93 |
| R\$ 3,001 – 10,000/month | 1.61 | 1.33-1.94 | 1.54 | 1.27-1.86 | 1.25 | 1.04-1.51 |
| Above R\$ 10,000/month | 1 | -- | 1 | -- | 1 | -- |
| *Change of family income* |  |  |  |  |  |  |
| Income did not drop | 1 | -- | 1 | -- | 1 | -- |
| Income dropped | 1.41 | 1.21-1.65 | 1.31 | 1.12-1.54 | 1.06 | 0.90-1.24 |
| *Provider of family income* |  |  |  |  |  |  |
| Responder is main provider | 1 | -- | 1 | -- | 1 | -- |
| Responder is not main provider | 1.34 | 1.13-1.59 | 1.35 | 1.13-1.60 | 0.99 | 0.83-1.17 |
| **D – PHYSICAL CONDITIONS, COMORBIDITIES, HABITS AND BEHAVIORS** |  |  |  |  |  |  |
| BMI |  |  |  |  |  |  |
| Normal | 1 | -- | 1 | -- | 1 | -- |
| Overweight/Obese | 1.05 | 0.90 – 1.23 | 1.01 | 0.86-1.17 | .98 | 0.84-1.14 |
| Comorbidities or other health conditions |  |  |  |  |  |  |
| No comorbidities | 1 | -- | 1 | -- | 1 | -- |
| With comorbidity(s) | 1.07 | 0.90-1.27 | 1.15 | 0.97-1.36 | 1.33 | 1.12-1.57 |
| Smoker |  |  |  |  |  |  |
| No | 1 | -- | 1 | -- | 1 | -- |
| Yes | 1.50 | 1.10-2.04 | 1.21 | 0.90-1.63 | 2.33 | 1.70-3.20 |
| Alcohol intake |  |  |  |  |  |  |
| No | 1 | -- | 1 | -- | 1 | -- |
| Yes | 0.96 | 0.83-1.13 | 0.73 | 0.63-0.85 | 1.15 | 0.97-1.34 |
| Symptoms of COVID-19 |  |  |  |  |  |  |
| No symptoms or COVID-19 was ruled out | 1 | -- | 1 | -- | 1 | -- |
| Had symptoms, COVID-19 diagnosed or tests not ready | 2.10 | 1.63-2.71 | 1.76 | 1.39-2.23 | 1.67 | 1.32-2.12 |
| Fear of contracting COVID-19 (excluded people who contracted) OBS: compared to “little worried” |  |  |  |  |  |  |
| Did not contract to the date |  |  |  |  |  |  |
| - Not or little worried | 1 | -- | 1 | -- | 1 | -- |
| - Moderately worried | 1.90 | 1.53-2.37 | 1.60 | 1.28-2.00 | 1.38 | 1.11-1.71 |
| - Very much worried | 5.24 | 4.08-6.71 | 3.31 | 2.60-4.22 | 2.16 | 1.70-2.72 |
| Time spent gathering news about COVID-19 on any media OBS: compared to not following news |  |  |  |  |  |  |
| Does not follow news | 1 | -- | 1 | -- | 1 | -- |
| Follows the news |  |  |  |  |  |  |
| - Up to one hour/day | 0.80 | 0.56-1.14 | 0.90 | 0.63-1.29 | 0.92 | 0.65-1.31 |
| - 1-3 hours/day | 1.26 | 0.87-1.83 | 1.31 | 0.90-1.89 | 1.21 | 0.83-1.74 |
| - More than 3 hours/day | 1.71 | 1.12-2.59 | 1.39 | 0.92-2.09 | 1.61 | 1.07-2.42 |
| Practice of Exercises OBS: All compared to “practiced before, still practices > 3 times/week |  |  |  |  |  |  |
| Did not practice before the pandemic |  |  |  |  |  |  |
| - Remains sedentary | 2.51 | 2.01-3.13 | 2.02 | 1.62-2.53 | 1.73 | 1.39-2.16 |
| - Started practicing | 2.00 | 1.46-2.73 | 1.77 | 1.29-2.41 | 1.98 | 1.45-2.71 |
| Practiced before the pandemic |  |  |  |  |  |  |
| - Is not practicing now | 2.25 | 1.80-2.80 | 2.39 | 1.91-2.99 | 1.50 | 1.20-1.87 |
| - Still practices, less than 3x/week | 1.36 | 1.06-1.75 | 1.09 | 0.84-1.41 | 1.30 | 1.01-1.67 |
| - Still practices, 3+ times/week | 1 | -- | 1 | -- | 1 | -- |
| Time preferred for practice of exercises (for those currently practicing) |  |  |  |  |  |  |
| Prefers morning | 1 | -- | 1 | -- | 1 | -- |
| Prefers afternoon | 1.39 | 1.04-1.85 | 1.42 | 1.05-1.91 | 1.38 | 1.03-1.85 |
| Prefers evening | 1.34 | 1.02-1.78 | 1.59 | 1.19-2.12 | 1.22 | 0.92-1.62 |
| Varies/no preference | 1.38 | 1.03-1.86 | 1.35 | 0.99-1.84 | 1.28 | 0.95-1.72 |
| Time spent on social activities (SA) |  |  |  |  |  |  |
| Less than before the pandemic | 1.28 | 1.05-1.56 | 1.62 | 1.34-1.97 | 1.66 | 1.37-2.02 |
| Equal/more than before the pandemic | 1 | -- | 1 | -- | 1 | -- |
| Time spent in instrumental activities (IA) |  |  |  |  |  |  |
| Less than before the pandemic | 1.67 | 1.27—2.19 | 1.63 | 1.26-2.11 | 1.75 | 1.35-2.28 |
| Equal/more than before the pandemic | 1 | -- | 1 | -- | 1 | -- |

**Table 4.**
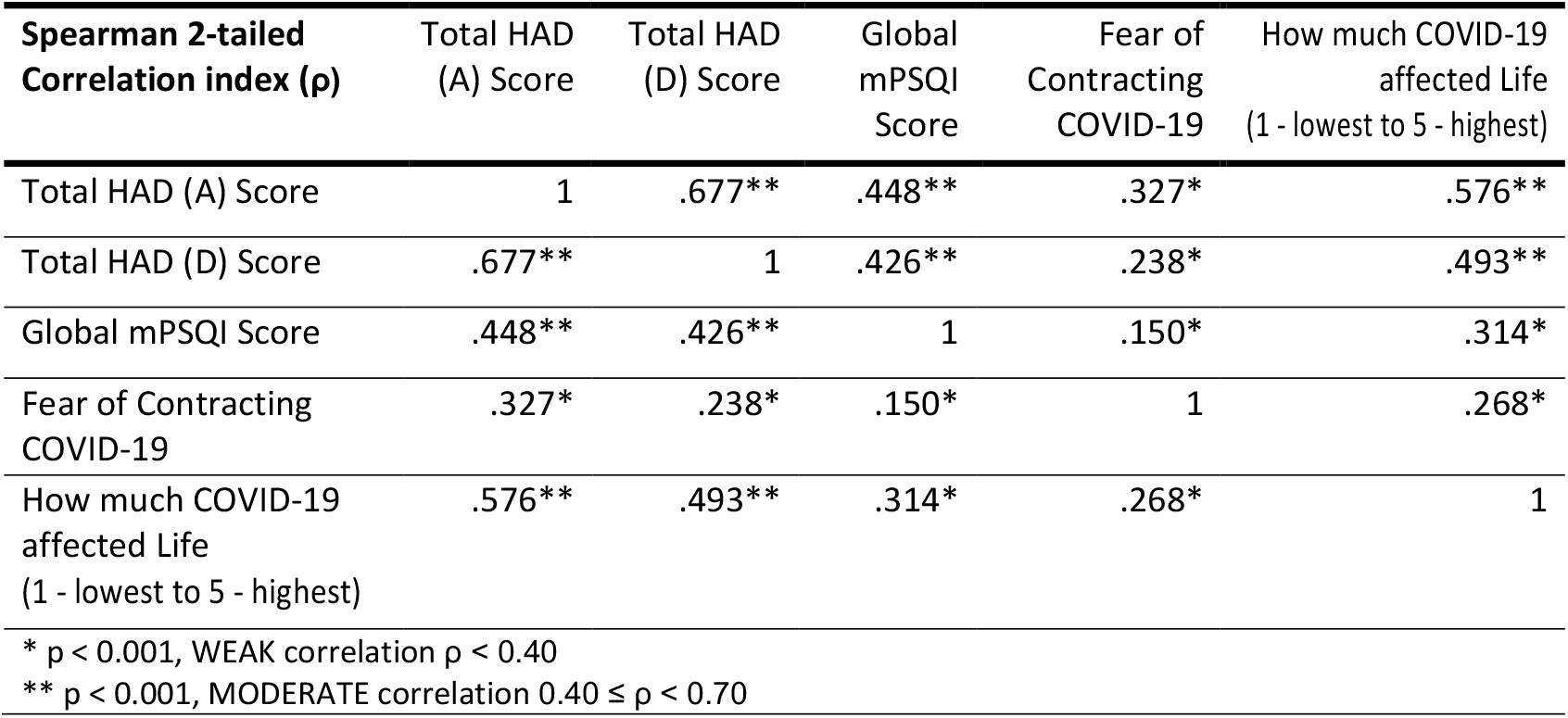
Two-tailed Spearman Correlation index among HAD (A) and (D) Scores, modified PSQI (mPSQI) Score, Fear of Contracting COVID-19 and Likert-scale grade (1 to 5) for How much COVID-19 was perceived to affect responder’s life, in general.

| **Spearman 2-tailed**<br>**Correlation index (ρ)** | **Total HAD**<br>**(A) Score** | **Total HAD**<br>**(D) Score** | **Global**<br>**mPSQI**<br>**Score** | **Fear of**<br>**Contracting**<br>**COVID-19** | **How much COVID-19**<br>**affected Life**<br>**(1 - lowest to 5 - highest)** |
| --- | --- | --- | --- | --- | --- |
| Total HAD (A) Score | 1 | .677** | .448** | .327* | .576** |
| Total HAD (D) Score | .677** | 1 | .426** | .238* | .493** |
| Global mPSQI Score | .448** | .426** | 1 | .150* | .314* |
| Fear of Contracting<br>COVID-19 | .327* | .238* | .150* | 1 | .268* |
| How much COVID-19<br>affected Life<br>(1 - lowest to 5 - highest) | .576** | .493** | .314* | .268* | 1 |

\* $p < 0.001$ , WEAK correlation $\rho < 0.40$
\*\* $p < 0.001$ , MODERATE correlation $0.40 \leq \rho < 0.70$

### III-B1. Demographics

Prevalence of mental and sleep disorders were higher in younger people, especially in the first quartile (ages 18 to 22). Meanwhile, the oldest quartile (ages 39 to 79) was statistically less anxious and depressed than expected.

Anxiety and depression were significantly higher in mestizos, and significantly lower in whites. There was no significance in differences of sleep quality among ethnicities.

There were no statistical disparities of mental or sleep issues when people who lived alone were compared to people living with more people during the pandemic.

### III-B2. Education and Occupation

People with less years of education tended to have more anxiety, depression and bad sleep symptoms, while those who had postgraduation degrees were significantly suffering less from those conditions.

Unemployed (regardless of being discharged more or less recently) and students had a higher prevalence of anxiety, depression and bad sleep quality than freelancers, workers of public or private sectors or those who were retired or received pension.

Rates of possible conditions were similar among non-health workers and health workers (regardless of dealing or not with COVID-19 cases).

Caregivers were significantly more prone to mental and sleep disorders than non-caregivers, especially depression.

### III-B3. Financial Status

Responders of families with lower monthly incomes and/or whose income had dropped during the pandemic were more prone for those conditions. Also, those who did not contribute to the family budget, or were not the main contributors were also more prone to possible anxiety and depression.

Sleep quality did not vary significantly among people with different family income, drop in family budget or among providers and non-providers.

### III-B4. Physical Conditions, Comorbidities, Habits and Behaviors

Weight did not seem to interfere in rates of disorders. Having or not comorbidities and being or not a smoker also did not interfere with anxiety and depression rates. People with no comorbidities were less prone to be bad sleepers than otherwise. Rates of bad sleep quality in smokers up to 10 cigarettes per day (65.6%) and in smokers of more than 10 cigarettes per day (73.3%) were significantly higher than in non-smokers (47.8%).

Rates of anxiety and bad sleep did not vary greatly among those who did not drink and alcohol consumers. The rate of depression was higher than expected in non-drinkers (49.6%), and lower than expected in those drinking up to twice a week and did not consume in the evening (41.6%). In general, alcohol drinkers were 27% less likely to be diagnosed with depression, in our sample. Nocturnal drinking did not play a statistically significant difference in rates.

Participants who did not had symptoms of COVID-19, by the time of the survey, were less likely to be anxious, depressed or bad sleepers than those with symptoms, regardless of confirmation of infection by the virus.

People who watched and/or read more than 3 hours per day of news were 71% (CI 1.12-2.59) more likely to be in the anxious group, and 61% (CI 1.07-2.42) more likely to be in the bad sleepers’ group, when compared to those who did not follow news. They were also 39% more likely to be in the depressed group, although in this case, the CI of 0.92-2.09 did not allow a concrete confirmation of this trend.

The three disturbances had the highest ratios in those who were exercising much less than before the pandemic and in those who were sedentary by the time of the collection of data, regardless of being active or not before the pandemics. For those who were active, responders that preferred exercising in the morning had the lowest ratios of anxiety, depression and bad sleep, although only depression reached statistical significance.

Lastly, patients were asked about the time spent in social activities (SA), like phoning or video-calling friends and relatives, engaging in social interactions and relationships, and in instrumental activities (IA), such as care of offspring, parents or other relatives, pet sitting, cleaning the house, religious activities. People who increased time spent both in SA and IA were significantly less prone to anxiety, depression and bad sleep. People who spent less time in SA than before COVID-19 were 28%, 62% and 66% more likely to be in the anxiety, depression and bad sleep groups, respectively, while those who spent less time in IA than before the infection were 67%, 63% and 75% more likely to be in the anxiety, depression and bad sleep groups, respectively.

### III-B5. Correlations

**Table 4** shows there was a moderate correlation between Anxiety Score – HAD (A) and Depression Score – HAD (D), and between Global mPSQI score and both HAD (A) and HAD (D). Grading of how much COVID-19 had affected the responder’s life (in a Likert scale, from 1 to 5) correlated moderately with both HAD (A) and HAD (D), but weakly with mPSQI. **Graph 1** depicts the cumulative dispersion of grading versus HAD (A), HAD (D) and mPSQI. Fear of contracting COVID-19 also displayed a weak correlation with other variables.

**Graph 1.**
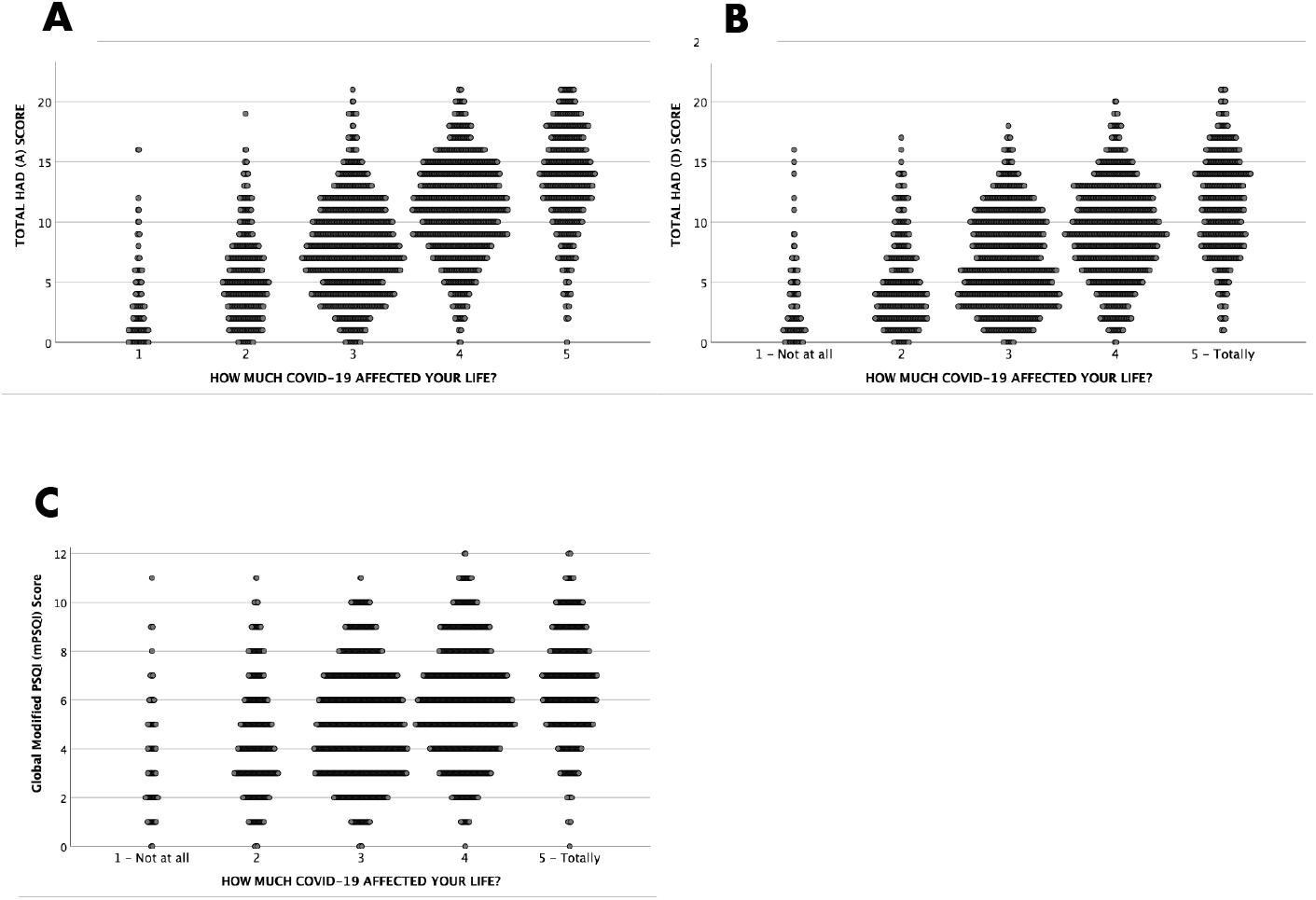
Cumulative dispersion of the Likert-scale grade for how much COVID-19 was perceived to affect responder’s life versus HAD (A) Score in **A**, HAD (D) Score in **B** and Global modified PSQI in **C**

## IV. Discussion

In this survey, we found global rates of 56.5% for possible anxiety, 46.1% for possible depression and 49.2% for bad sleep quality.

A meta-analysis conducted by Salari et. al (28) found, among 17 studies conducted in 2020, anxiety rates varying between 6.33% and 50.9% (mean of 32.9% for Asian and 23.8% for European studies) and depression rates from 17.17% to 53.5% (mean of 35.3% for Asian and 32.4% for European studies). Assessment for those conditions was heterogeneous. The most frequent used instruments were the DASS-21 - Depression, Anxiety and Stress Scale (29), the GAD-7 – Generalized Anxiety Disorder 7-item (30), and the PHQ-9 – Patient Health Questionnaire (31).

Rates of anxiety and depression in our survey were higher than the averages of the aforementioned meta-analysis, however some factors must be bore in mind: first, none of those studies used the HAD scale for anxiety and depression, adopted in this study. Secondly, and in particular regarding anxiety rate, studies that used the DASS-21 scale also assessed stress levels. Some of the stress questions are of resemblance with anxiety questions of the HAD scale - for instance: “I found it hard to wind down” is accounted for stress in DASS-21, and “I feel tense or ‘wound up’” is accounted for anxiety in HAD. This raises a possibility that, in studies based on the DASS-21, anxiety rates were lower due to a split caused by the stress group. Thirdly, none of those studies included were from Latin-American countries. Another Brazilian study (32), with 45,161 participants, intended to evaluate sadness/depression, anxiety/nervousness and sleep problems. Data was collected online between April 24^th^ and May 24^th^, 2020, a time window very much similar to ours. Although using a much simpler form, consisting of a few questions – i.e., not using standardized, validated questionnaires, this survey found rates of 52.6% of anxiety/nervousness, 43.5% of sadness/depression and 43.5% of an onset of sleep issues, percentages that resemble ours.

Studies conducted out of a pandemic scenario shows lower rates of the same disturbances. A Brazilian, multicenter survey of 2014 (33), was conducted with data from 1,857 participants who were assessed when using primary healthcare, from four Brazilian cities. This study also used the HAD scale as instrument to access mental disorders. Anxiety rates oscillated from 35.4% to 43%, and depression rates ranged between 21.4% and 31%. In our survey, anxiety and depression rates (see **Tables 2a-d**) are higher. Another study (34) estimated, by analysis of 12,000 people that responded to the 2011 National Health and Wellness Survey in Brazil, the prevalence of Major Depression to be 10.2%, again much lower than what we found.

Our data shows that youngers, females, students, people with lower familiar monthly income or a significant drop of income during the epidemic, unemployed, people following more hours per day of news concerning COVID-19, people engaging less in exercising and in social and instrumental activities were groups associated with greater odds of being possibly anxious, depressed and/or bad sleeper. Younger age and more time following news were also associated with higher levels of mental and sleep issues in a Chinese survey of 7,236 volunteers, conducted by Huang and Zhao (8), corroborating our findings for these groups.

Mestizos were more likely to be possibly depressed. Although rates of one or both disturbances were also higher in blacks and native Brazilians (indigenous), this did not reach statistical significance, possibly due to low sampling. The disproportional high rate of whites (74.1%) and low rates of mestizos (19.4%), blacks (4.4%) and other ethnicities responding (latest Brazilian census shows 47.5% of auto-declared whites, 43.2% of mestizos and 7.5% of blacks (35)) are clear evidence of the social and economic inequality in our local reality (36).

BMI did not influence rates of anxiety, depression and bad sleep, while the absence of comorbidities was a protective factor for better sleep quality, but not for anxiety and depression. Sedentary people (regardless of being or not active before the pandemic) were more prone all those conditions, and those who already practiced before and kept practicing more than 3 times a week showed the least tendency to mental and sleep disturbances, showing that exercising played a role in preventing worsening of mental and sleep status in our sample. Also, of note, people that preferred to exercise early were less depressed. That could be due a bidirectional relationship, as depression is commonly linked with morning fatigue and unwillingness to engage in energetic activities (37).

Curiously enough, those who did not consume alcohol had a significant higher rate of depression than alcohol consumers. We do not propose a logical explanation, but researchers should pay attention to forthcoming articles, to see if this pattern replicates, or if it could be merely coincidental. Conversely, despite not reaching significant differences, but towards what is traditionally more expected, alcohol consumers, especially those with nocturnal consumption, had higher rates of anxiety and bad sleep quality. Smokers had greater incidence of bad sleepy quality, especially those who smoked more than 10 cigarettes per day. This is expected, as smoking is known to be disruptive to good sleep (38).

Also relevant is the observation that, in most of the similar studies we found so far, where snowball recruiting was performed - therefore sampling was not randomized but rather by convenience, younger adults (up to the fourth decade of life) and females were the most solicitous. Female participation was majoritarian in most studies conducted worldwide, with encountered proportions of 51% (12) 53.8% (17), 54.6% (8), 64.7% (15), 66% (10), 76.1% (16), 76.3% (our data) and 81.1% (9). Females were the minority in two studies, with proportions of 41.6% (11) and 45.5% (13). Mean ages, for papers that provided this asset, were of 31.7 (our data), 33 (11), and 35.3 (8). Other papers provided age ranges: one study disclosed that 56.5% of participants were from 18 to 25 years-old (9), in another 77% of participants were from 18 to 40 years-old (10), and yet in other 47.3% were aged between 18-35 years, and an additional 33.8% were from 36 to 45 years-old (17). While youth is naturally expected to outweigh middle agers and seniors, due to easiness of interaction with digital technology, the more participation of females could be multifactorial.

Apart from snowball sampling, there were other limitations. By lowering by 7 points the maximum punctuation possible of the PSQI, the authors acknowledge that sensitivity might have been diminished for detection of bad sleepers, but a necessary cost, given the facts already explained in methodology. Nonetheless, we consider that the specificity of the diagnosis of bad sleep remained unharmed, and the high proportion of bad sleepers, with the mPSQI, is by itself a fact worthy of consideration.

## V. Conclusion

The DEGAS-CoV study, in accordance to many published papers so far, shows high rates of anxiety (56.5%), depression (46.1%) and bad sleep quality (49.2%) during COVID-19. These rates reflect the time of data collection, in the second trimester of 2020, in the Brazilian population. Some groups were more prone to those disturbances, such as: younger people, women, mestizos, people with lesser years of education, of lower income or whose income dropped significantly during the pandemic, unemployed, caregivers, people who followed more hours of news of COVID-19, sedentary or those who engaged less in physical activity, and those less engaged in social and instrumental activities. More studies, especially longitudinal, could provide more information on if the prevalence of mental and sleep issues remains constant during the pandemic crisis or suffer changes related to different aspects, like adaptation to the situation, social restriction relaxation and improved treatments or vaccines. We also intend to conduct further sub-analyses of our data, in search of more significant statistical findings.

## Data Availability

Data available within the article or its supplementary materials

